# Body Mass Index and Non-invasive Cardiovascular Parameters

**DOI:** 10.1101/2024.05.17.24307560

**Authors:** Shenghui Wu, Marco Meucci, Zhong Liu

**Affiliations:** Department of Public Health and Exercise Science, Beaver College of Health Sciences, Appalachian State University, 1179 State Farm Road, Boone, NC, 28607

**Author notes:** Corresponding and senior author: Shenghui Wu, MD, PhD, Department of Public Health and Exercise Science Beaver College of Health Sciences, Appalachian State University Boone, NC 28607.

**Keywords:** body mass index, cardiovascular health, pulse wave velocity, AIx75, ejection time, Buckberg

## Abstract

**Background:** Epidemiological studies on body mass index (BMI) and non-invasive cardiovascular parameters are limited and inconsistent. To provide more informative data for further prevention and control, we examined associations between BMI, as well as overweight/obesity, and non-invasive cardiovascular parameters and their dose-response relationships in North Carolina Appalachian adults.

**Methods:** A total of 71 participants were included in this analysis. Non-invasive cardiovascular parameters included carotid-femoral pulse wave velocity for measuring central arterial stiffness, augmentation index at 75 bpm for gauging peripheral arterial stiffness, ejection time for indicating left ventricular performance, and Buckberg index for measuring coronary microvascular circulation. Logistic regression models were used for analysis.

**Results:** Every unit (kg/m^2^) increase in BMI was associated with a 25% statistically significant increased multivariable-adjusted odds of higher central arterial stiffness (odds ratio: 1.25; 95% confidence interval: 1.04-1.51), a 31% increased adjusted odds of higher peripheral arterial stiffness, a 23% statistically significant increased adjusted odds of worse left ventricular performance, and a 25% statistically significant increased adjusted odds of worse coronary microvascular circulation. Overweight/obesity was associated with a 532% statistically significant increased odds of higher arterial stiffness (6.32; 1.42-28.09), and a 704% statistically significant increased odds of worse left ventricular performance after adjusting for age, sex, physical activity, and body fat percentage.

**Conclusion:** Increased BMI, especially overweight/obesity, was significantly associated with the increased risk of worse cardiovascular health, measured by non-invasive cardiovascular parameters. Efforts need to be focused on improving interventions to lower BMI/reduce overweight and obesity in North Carolina, especially in Appalachian populations.

## Introduction

Cardiovascular disease (CVD) is the first leading cause of death in the world^1^ and the US^2^. High body mass index (BMI), as a preventable behavior, poses a heavy burden on the US healthcare system^3,4^, and was suggested to be positively associated with CVD risk factors and outomes^5,6^. Non-invasive cardiovascular parameters are very practical and feasible in population-based studies due to their easy and non-invasive operations. However, epidemiological studies on BMI and non-invasive cardiovascular parameters are limited, provided only correlations without consideration of confounding factors, and showed inconsistent results. For example, pulse wave velocity (PWV) reflects segmental arterial elasticity, and it is an indicator of arterial stiffness. The pulse wave created by ventricular propagates throughout the arterial tree at a certain velocity that depends on the pressure gradient, the diameter of the artery and the total peripheral resistances. Increase of the arterial stiffness is associated with increase of the propagation speed of the pulse wave in the arteries ^7–9^. PWV was positively correlated with BMI in school-aged children (*r* = 0.237, *p* < 0.05) ^10^ and adults (*r* = 0.206, *p* < 0.05)^11^. Carotid-femoral PWV (cfPWV), calculated as the distance traveled by the pulse wave from carotid to femoral artery divided by the transient time, it is a strong independent predictor of CVD risk in both general and patient populations^7–9^. It advised the use of ≤10 m/s as cut-off value for cfPWV^8,9^. Augmentation index at 75 bpm (AIx75) is a measure of peripheral arterial stiffness. The stiffer the peripheral arteries are, the earlier and augmented will be the reflection of the pulse wave from the periphery ^7,12–14^.

AIx75 was positively correlated with body fat mass (*r* = 0.154, *p* < 0.05), but negatively correlated with BMI (*r* = -0.209, *p* < 0.05) in school-aged children, with stronger correlations in students with obesity (*r* = 0.573 and *r* = -0.410, respectively)^10^. Higher arterial stiffness augments systolic pressure and consequently AIx which represents the afterload inflicting the ventricles. Elevated arterial stiffness increases the risk of development of cardiovascular events and impairs cardiovascular functions^7,12–14^. While no threshold values have been described in the literature for AIx75, it has been suggested that 27.6% is an appropriate cutoff for AIx75 (SphygmoCor) ^14^. Ejection time (ET), also known as Left Ventricular Ejection Time (LVET), is the time interval from aortic valve opening to aortic valve closure. It is the phase of systole during which the left ventricle ejects blood into the aorta. ET has been used for several decades to assess left ventricular function and contractility.

For example, ET is reduced in patients with reduced ejection fraction ^15–18^. It was advised that the cutoff seems to be below 350 ms^18^. Finally, the Buckberg index, also known as Subendocardial Viability Ratio is an index of myocardial oxygen supply and demand^19^. SEVR is also an indicator of arterial stiffness correlated with coronary flow reserve, which makes it a useful parameter in assessing coronary microvascular circulation, especially in hypertensive patients^20,21^. A cutoff of 130% was suggested ^22^.

To provide more informative epidemiological data for further prevention and control, we examined associations between BMI, as well as overweight/obesity, and non-invasive cardiovascular parameters and their dose-response relationships.

## Methods

### Participants

This study received approval from the Appalachian State University Institutional Review Board review (IRB# 21-0167). Participants were recruited by word of mouth, flyers, and Appalachian State University website. Individuals from the state of North Carolina participated in this study. Participants were Appalachian State University students and community members. The majority of the participants were from Watauga County. Participants were included in this research if they were between 18 and 65 years of age, and they were excluded if they were diagnosed with cardiovascular, renal and metabolic diseases or if they had related symptoms, and if they were taking medications associated with those conditions. Participants gave written consent to participate in the research. Exclusion criteria included medical conditions such as diabetes, heart, respiratory, or renal disease, and taking any medications at the study onset. Participants were instructed to report to the laboratory after a minimum 4-hour abstention from caffeinated drinks, alcohol and nicotine, and large meals. Also, participants were asked not to exercise on the day of the test. Measurements were performed between 2:00 to 6:00 PM. Data have been collected from 2018 and 2023. Demographic information included age, sex, race, and ethnicity. Physical activity including walking and other exercises in the last week was collected.

### Anthropometrics and Body Composition

Height and body mass were measured with a stadiometer and a scale to the nearest 0.1 cm and 0.1 kg, respectively. BMI was calculated using body weight and height (BMI = (kg/m^2^)). Fat mass (FM) in absolute terms and relative to total body weight (FM%) were assessed in all subjects via an air displacement plethysmography (BodPod gold, COSMED, Italy) using the Siri equation and predicted thoracic gas volume or a Horizon® Dual-energy X-ray absorptiometry System (DEXA, Horizon, Hologic, Marlborough, MA) with participants in a supine position with feet internally rotated and hands flat on the test table. DEXA was the preferred assessment, body composition was assessed through BodPod if participants did not receive permission from their physician to perform a DEXA scan, if they were pregnant, if they had metal implants in their body, or if they had a body weight grater than 450 lb. Subjects wore tight-fitting clothes, a swimmer’s cap, and no shoes or jewelry during the body composition assessment. Participants were categorized as normal weight (<25 kg/m^2^), overweight (25–29.9 kg/m^2^), or obese (≥30 kg/m^2^) using BMI cutoffs of 25.0 and 30 kg/m^2^ ^23^.

### Cardiovascular measurement

Blood pressure and arterial stiffness were assessed with an automated SphygmoCor XCEL device (SphygmoCor, AtCor Medical, Inc.). PWA and cfPWA were assessed in a dimly lit room, after 5 minutes of quiet rest sitting, and lying supine, respectively. Brachial blood pressure was taken from the right arm and the PWA output provided by the instrument included aortic AIx normalized to 75 bpm (AIx75), ET, and Buckberg index. cfPWV was calculated as the quotient of the vascular distance in meters and the pulse wave transit time in seconds (m/s). The vascular distance was determined by the SphygmoCor XCEL software using the distance between the carotid artery to the sternal notch, the sternal notch to the leg cuff, and the femoral artery to the leg cuff. Transit time was measured using volumetric displacement (femoral artery) and applanation tonometry (carotid artery), with all distances measured using a fabric tape by a trained technician. Recordings of the waveforms with consistent amplitude and shape that resulted in a quality control index equal to or greater than 90% were used for statistical analysis. All measurements were performed for each subject a minimum of three times for PWA and twice for cfPWV with one-minute rest intervals. The two closest trials for each participant with systolic and diastolic pressure within ±5 mmHg for PWA ^24^ and within ± 0.3 m/s for PWV ^25^ were averaged and used for the statistical analysis.

### Statistical analysis

Chi-square tests for categorical variables and Student t-tests/Wilcoxon rank-sum tests for continuous variables were conducted to assess differences between groups as appropriate. Logistic regression models were used to estimate the odds ratios (ORs) and their 95% CIs for associations between BMI or overweight/obese and cardiovascular parameters after adjusting for other covariates. cfPWV was divided into two groups according to the median of cfPWV (high arterial stiffness: ≥6.45 m/s vs. low arterial stiffness <6.45 m/s) since all participants had cfPWV ≤10ms which is the suggested cut-off point ^8,9^. AIx75 was classified into two groups based on the cut-off value of 27.6% (high peripheral arterial stiffness: ≥27.6% vs. low peripheral arterial stiffness: <27.6%)^14^. Ejection time was separated into two groups as per the median of ET (worse left ventricular performance: <299.5 ms vs. better left ventricular performance ≥299.5 ms) since all but one participant had ET ≥350 ms which is the advised cut-off value ^18^. Buckberg was classified into two groups based on the median (worse coronary microvascular circulation: <181% vs. better coronary microvascular circulation: ≥181%) because all but two participants had Bucberg index values ≤130% which is the suggested cutoff point ^22^. Covariates included age, sex, physical activity, and body fat percentage. All participants were nonsmokers, all but three were White, and all but two were non-Hispanics, therefore, smoking status, race, and ethnicity were not adjusted for in regression models. We also used a restricted cubic spline logistic regression analysis to evaluate the association of ORs of cardiovascular parameters with BMI on a continuous basis after adjusting for other covariates. Tests of statistical significance were based on two-sided probability, and *P* < 0.05 was considered statistically significant. Statistical modeling was performed by using SAS 9.4 (SAS Institute, Cary, NC).

## Results

A total of 71 adults with available data were included in this analysis. The mean age was 45.9 (standard deviation: 15.7) years; and 45% were males (**Table 1**). A total of 45% of the participants were overweight or obese. Compared with those with normal weight, participants with overweight/obesity were more likely to have higher body fat percentage, higher levels of PWV, and lower levels of ejection time (all *P*s < 0.05).

**Table 1.**
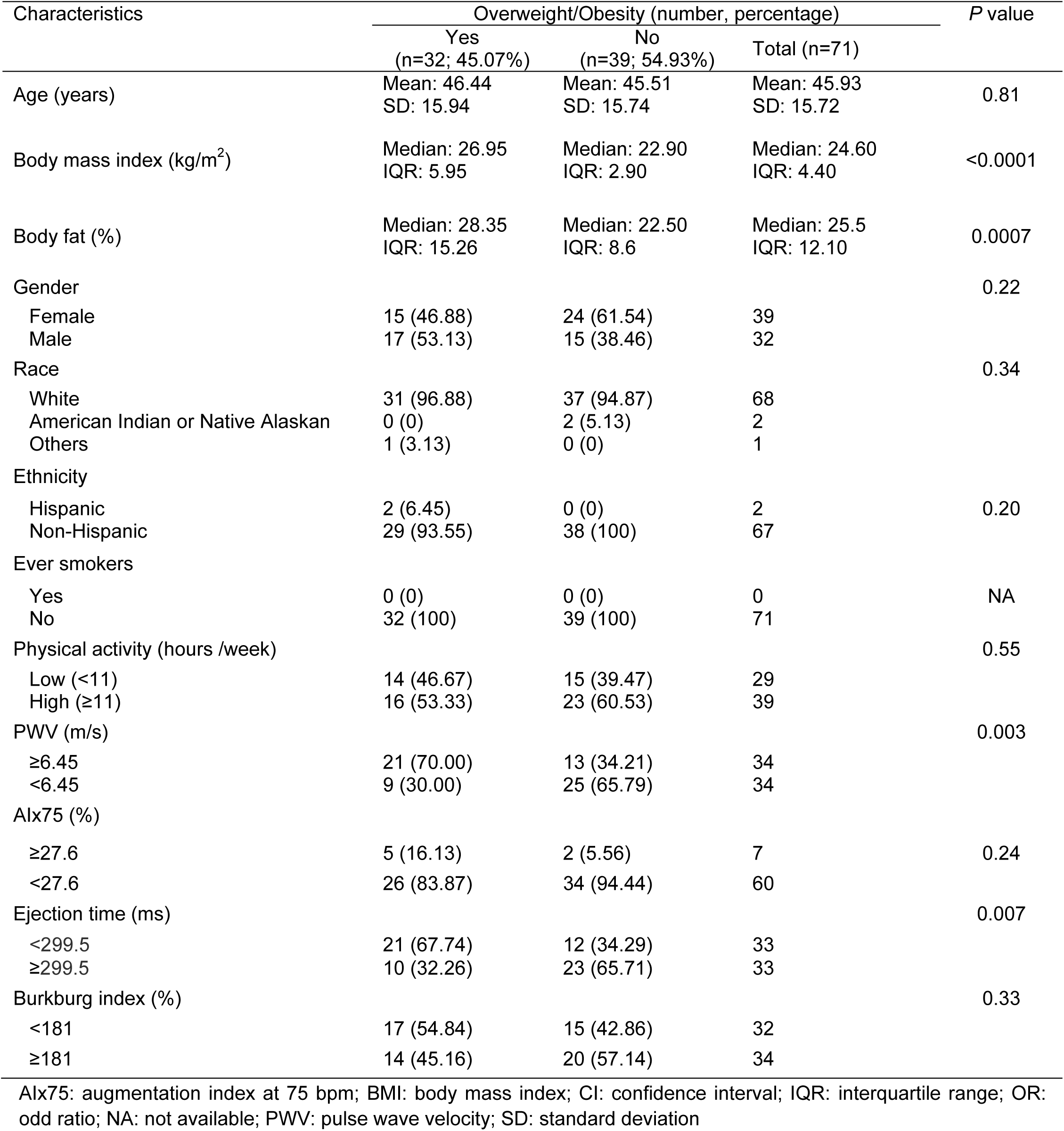
Characteristics Stratified by Overweigt/Obesity Status.

The statistically significant positive associations were observed between BMI and PWV as well as between overweight/obesity and PWV for both crude and adjusted models. Every unit (kg/m^2^) increase in BMI was associated with 25% statistically significant increased adjusted odds of a higher level of PWV (high arterial stiffness: ≥6.45 m/s vs. low arterial stiffness <6.45 m/s). Overweight/obesity was associated with a 532% (OR: 6.32; 95% CI: 1.42-28.09) statistically significant increased adjusted odds of a higher level of PWV (**Table 2**).

**Table 2.** Logistic Regression Analyses for The Associations Between Body Mass Index and Cardiovascular Health Indicators.

| Cardiovascular Health Indicators | Crude OR (95% CI) | *P* | Adjusted OR (95% CI)<sup>a</sup> | *P* |
| --- | --- | --- | --- | --- |
| **PWV (≥6.45 m/s vs. <6.45 m/s)** |  |  |  |  |
| BMI (kg/m<sup>2</sup>) | 1.18 (1.03-1.35) | 0.02 | 1.25 (1.04-1.51) | 0.02 |
| Obesity |  |  |  |  |
| Normal (BMI<25 kg/m<sup>2</sup>) | Reference |  | Reference |  |
| Overweight/obesity (BMI≥25 kg/m<sup>2</sup>) | 4.49 (1.60-12.56) | 0.004 | 6.32 (1.42-28.09) | 0.02 |
| **AIx75 (≥27.6% vs. <27.6%)** |  |  |  |  |
| BMI (kg/m<sup>2</sup>) | 1.17 (1.05-1.30) | 0.005 | 1.31 (0.99-1.73) | 0.06 |
| Obesity |  |  |  |  |
| Normal (BMI<25 kg/m<sup>2</sup>) | Reference |  | Reference |  |
| Overweight/obesity (BMI≥25 kg/m<sup>2</sup>) | 3.27 (0.59-18.21) | 0.18 | 1.90 (0.21-17.19) | 0.57 |
| **Ejection time (< 299.5 ms vs. ≥299.5 ms)** |  |  |  |  |
| BMI (kg/m<sup>2</sup>) | 1.10 (0.99-1.21) | 0.07 | 1.23 (1.04-1.45) | 0.01 |
| Obesity |  |  |  |  |
| Normal (BMI<25 kg/m<sup>2</sup>) | Reference |  | Reference |  |
| Overweight/obesity (BMI≥25 kg/m<sup>2</sup>) | 4.02 (1.44-11.24) | 0.008 | 8.04 (2.09-30.89) | 0.002 |
| **Buckberg index (<181% vs. ≥181%)** |  |  |  |  |
| BMI (kg/m<sup>2</sup>) | 1.17 (1.03-1.33) | 0.01 | 1.25 (1.06-1.48) | 0.01 |
| Obesity |  |  |  |  |
| Normal (BMI<25 kg/m<sup>2</sup>) | Reference |  | Reference |  |
| Overweight/obesity (BMI≥25 kg/m<sup>2</sup>) | 1.62 (0.61-4.29) | 0.33 | 1.05 (0.33-3.35) | 0.94 |

AIx75: augmentation index at 75 bpm; BMI: body mass index; CI: confidence interval; OR: odd ratio; PWV: pulse wave velocity
<sup>a</sup>Adjusted for age, sex, physical activity, and body fat percentage.

BMI was positively associated with AIx75 in the crude model, and the positive association became borderline significant after adjusting for sex, age, physical activity, and body fat percentage of diagnosis (**Table 2**). Every unit (kg/m^2^) increase in BMI was associated with 31% increased odds of a higher level of AIx75 (high peripheral arterial stiffness: ≥27.6% vs. low peripheral arterial stiffness: <27.6%). Although the association between overweight/obesity and AIx75 was not statistically significant, their potential positive direction was indicated.

The statistically significant positive associations were observed between BMI and ejection time as well as between overweight/obesity and ejection time for both crude and adjusted models. Every unit (kg/m^2^) increase in BMI was associated with 23% statistically significant increased adjusted odds of a lower level of ejection time (worse left ventricular performance: <299.5 ms vs. better left ventricular performance ≥299.5 ms). Overweight/obesity was associated with a 704% statistically significant increased adjusted odds of a lower level of ejection time (**Table 2**).

BMI was statistically significant and positively associated with the Buckberg index in both crude and adjusted models. Every unit (kg/m^2^) increase in BMI was associated with 25% increased odds of a lower level of the Buckberg index (worse coronary microvascular circulation: <181% vs. better coronary microvascular circulation: ≥181%). The association between overweight/obesity and the Buckberg index was not statistically significant, but the potential positive direction was suggested (**Table 2**).

Figure 1 visually depicts the dose-response association between BMI and ln(OR)s of worse cardiovascular health after adjusting for all potential confounding variables in a restricted cubic spline logistic regression model. BMI was statistically and positively associated with the lnOR of a higher level of PWV (*P* for overall relation = 0.04 and *P* for linear relation = 0.03) (Figure 1A). Every unit (kg/m^2^) increase in BMI was associated with 50% increased odds (OR: 1.50, 95% CI: 1.05-1.20) of a higher level of arterial stiffness (PWV: ≥6.45 m/s vs. <6.45 m/s). Although BMI was not statistically associated with the lnOR of a higher level of AIx75 (*P* for overall relation = 0.18 and *P* for linear relation = 0.55) (Figure 1B), and a lower level of ejection time (*P* for overall relation = 0.03 and *P* for linear relation = 0.07) (Figure 1C) as well as the Buckberg index (*P* for overall relation = 0.06 and *P* for linear relation = 0.49) (Figure 1D), the potential positive associations were suggested in these figures.

**Figure 1.**
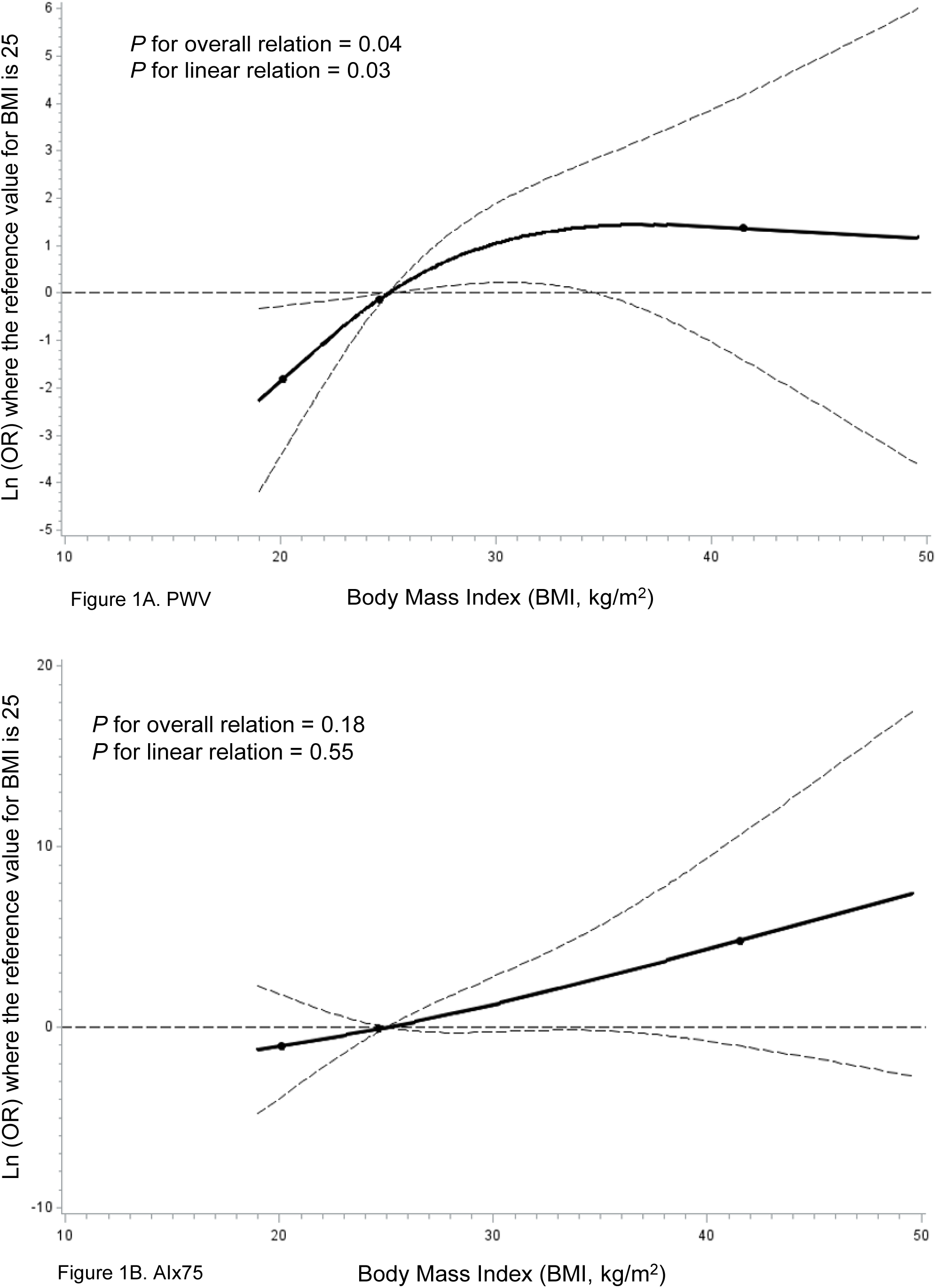
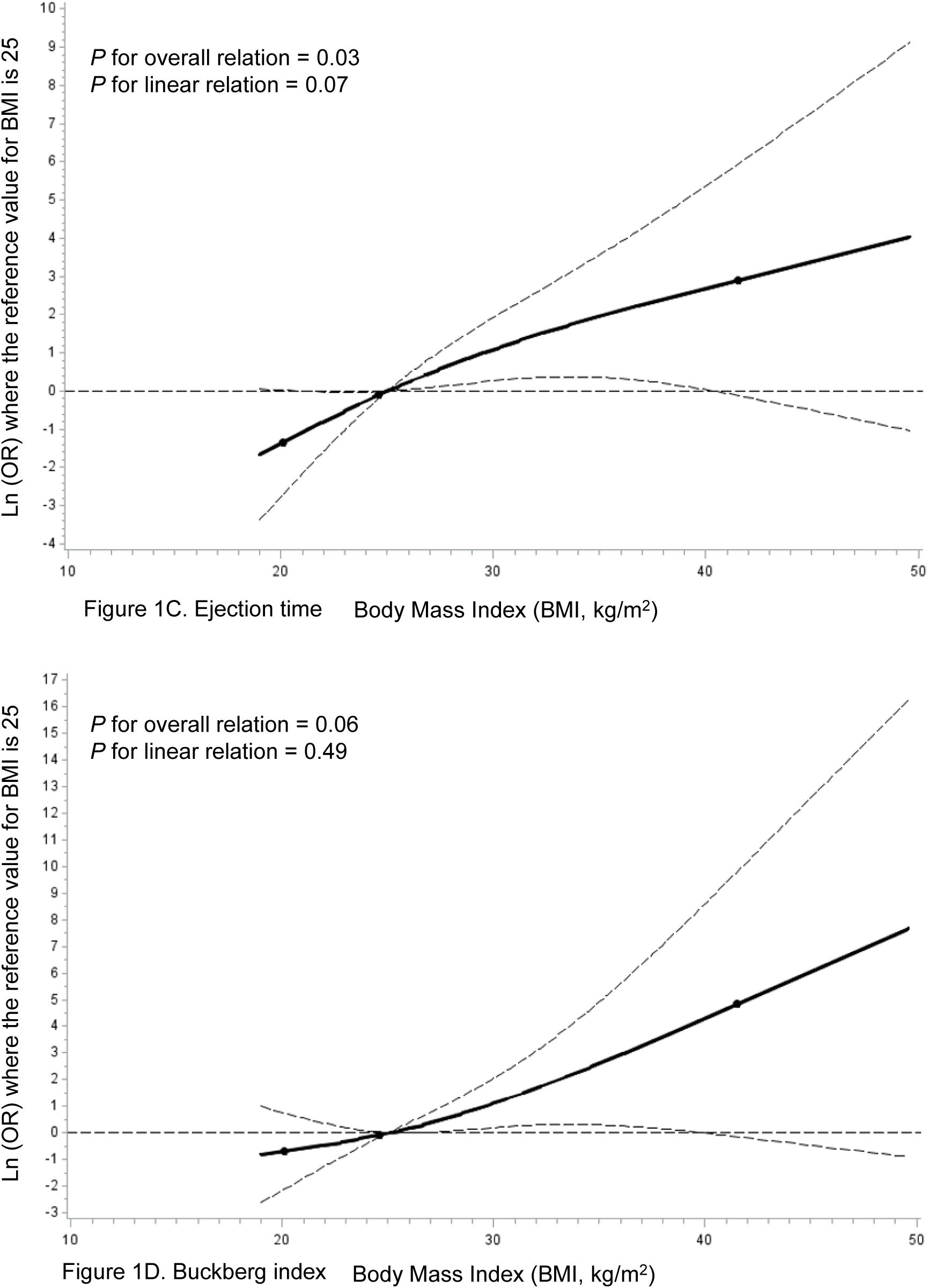
Adjusted Dose-response Association between Body Mass Index and Cardiovascular Health Indicators. Body mass index (BMI, kg/m^2^) was coded using a restricted cubic spline function with three knots located at the 5^th^, 50^th^, and 95^th^ percentiles of BMI distribution. *Y*-axis represents the adjusted ln(odds ratio) for worse cardiovascular health for any value of BMI compared to those with 25 of BMI adjusting for age, sex, race, physical activity, and body fat percentage. Dashed lines are 95% confidence intervals. Knots are represented by dots. Figure 1A: pulse wave velocity (PWV); Figure 1B: augmentation index at 75 bpm (AIx75); Figure 1C: Ejection time; Figure 1D: Buckberg index.

## Discussion

In a North Carolina Appalachian population sample, this study found that every unit (kg/m^2^) increase in BMI was associated with 25% statistically significant increased multivariable-adjusted odds of a higher level of PWV, 31% increased adjusted odds of a higher level of AIx75, 23% statistically significant increased adjusted odds of a lower level of ejection time, and 25% increased odds of a lower level of Buckberg index. Overweight/obesity was associated with 532% statistically significant increased odds of a higher level of PWV, and 704% statistically significant increased odds of a lower level of ejection time after adjusting for other potential confounding factors. These data strongly indicate the potential for BMI to have a profound effect on cardiovascular health which is measured by non-invasive cardiovascular parameters, and this is more evident because there appears to be a dose-response effect with higher BMI associated with even higher arterial stiffness.

Our study indicated that BMI was associated with increases in arterial stiffness (measured by PWV), peripheral arterial stiffness (measured by AIx75), worse left ventricular performance (measured by ejection time), and worse coronary microvascular circulation (measured by the Buckberg index). This suggests that the significant burden associated with overweight and obesity in North Carolina, especially among Watauga County residents, should be a focus. Our findings indicate that maintaining a healthy body weight, especially reducing BMI to be in the normal range, would be beneficial to reduce cardiovascular risk. Our findings were consistent with prior limited studies^10,11^. Several potential mechanisms might be involved in the role of obesity in the above cardiovascular problems. The development of arterial stiffness in obesity might be associated with stiffness of endothelial^26,27^ and vascular smooth muscle cells^28^, extracellular matrix remodeling^27^, perivascular adipose tissue inflammation^29^, and immune cell dysfunction^27^. Endothelial generation of nitric oxide is reduced by increased endothelial cortical stiffness^30,31^, and destruction of nitric oxide is promoted by enhanced oxidative stress^32^. Throughout the development of obesity, the adapted behavior of coronary resistance arteries leads to an inconsistency between blood supply and augmented metabolic requirement, which causes coronary microvascular disease^33^. Elevated aldosterone and vascular mineralocorticoid receptor activation might be involved in the development of cardiovascular stiffness^27^.

Our study has some limitations. First, the study used a cross-sectional design; thus, we could only examine an association but not a causal relationship. Second, BMI and cardiovascular parameter levels only reflected the participants’ BMI and cardiovascular parameter status when data were collected; thus, we could not know the temporal associations between BMI and cardiovascular parameters. The mechanisms and even dose-response of BMI on cardiovascular parameters could be further investigated in prospective cohort studies or randomized clinical trials. Third, we could not exclude the possibility of residual confounding due to unmeasured or inadequately measured covariates, but we controlled for available confounding variables in analyses. Last, dietary intake and other potential confounding factors were not collected in this study. However, future prospective cohort studies with a larger sample size will provide us with opportunities to include those factors.

Our research also had several strengths. To our knowledge, this is the first epidemiological study to examine associations between BMI and non-invasive cardiovascular parameters in North Carolina. Specifically, we identified the dose-response relationships between BMI and cardiovascular parameters, especially cfPWV, AIx75, ejection time, and the Buckberg index; these findings provided data for future research related to cardiovascular prevention and control. Finally, we included all available potential confounding factors, which allows us to get a relatively comprehensive analysis of the relevant factors.

In conclusion, increased BMI, especially overweight/obesity, was significantly associated with the increased risk of worse cardiovascular health, measured by non-invasive cardiovascular parameters, after excluding the effect of other confounding factors. Therefore, BMI might be a modifiable risk factor for which people can make changes to reduce their cardiovascular risk. Efforts need to be focused on improving interventions to lower BMI/reduce overweight and obesity in North Carolina, especially in Appalachian populations.

## Data Availability

The data that support the findings of this study are available on request from the author MM. The data are not publicly available due to privacy or ethical restrictions.

## References

1. World Health Organization. Cardiovascular diseases (CVDs). [Fact sheet]. 2021.

2. Centers for Disease Control and Prevention. Heart Disease Facts. https://www.cdc.gov/heartdisease/facts.htm. 2024. Accessed March 26, 2024.

3. Tsai AG, Williamson DF, Glick HA. Direct medical cost of overweight and obesity in the USA: a quantitative systematic review. Obes Rev. 2011;12:50–61. doi: 10.1111/j.1467-789X.2009.00708.x

4. Danaei G, Ding EL, Mozaffarian D, Taylor B, Rehm J, Murray CJ, Ezzati M. The preventable causes of death in the United States: comparative risk assessment of dietary, lifestyle, and metabolic risk factors. PLoS Med. 2009;6:e1000058. doi: 10.1371/journal.pmed.1000058

5. Kibret KT, Strugnell C, Backholer K, Peeters A, Tegegne TK, Nichols M. Life-course trajectories of body mass index and cardiovascular disease risks and health outcomes in adulthood: Systematic review and meta-analysis. Obes Rev. 2024;25:e13695. doi: 10.1111/obr.13695

6. McGee DL, Diverse Populations C. Body mass index and mortality: a meta-analysis based on person-level data from twenty-six observational studies. Ann Epidemiol. 2005;15:87–97. doi: 10.1016/j.annepidem.2004.05.012

7. Anastasio F, Testa M, Ferreri C, Rossi A, Ruocco G, Feola M. The Analysis of Arterial Stiffness in Heart Failure Patients: The Prognostic Role of Pulse Wave Velocity, Augmentation Index and Stiffness Index. J Clin Med. 2022;11. doi: 10.3390/jcm11123507

8. Van Bortel LM, Laurent S, Boutouyrie P, Chowienczyk P, Cruickshank JK, De Backer T, Filipovsky J, Huybrechts S, Mattace-Raso FU, Protogerou AD, et al. Expert consensus document on the measurement of aortic stiffness in daily practice using carotid-femoral pulse wave velocity. J Hypertens. 2012;30:445–448. doi: 10.1097/HJH.0b013e32834fa8b0

9. Mancia G, Fagard R, Narkiewicz K, Redon J, Zanchetti A, Bohm M, Christiaens T, Cifkova R, De Backer G, Dominiczak A, et al. 2013 ESH/ESC guidelines for the management of arterial hypertension: the Task Force for the Management of Arterial Hypertension of the European Society of Hypertension (ESH) and of the European Society of Cardiology (ESC). Eur Heart J. 2013;34:2159–2219. doi: 10.1093/eurheartj/eht151

10. He W, Zhang Y, Li X, Mu K, Dou Y, Ye Y, Liu F, Yan W. Multiple non-invasive peripheral vascular function parameters with obesity and cardiometabolic risk indicators in school-aged children. BMC Pediatr. 2022;22:146. doi: 10.1186/s12887-022-03214-4

11. Zuo J, Tang B, O’Rourke MF, Avolio AP, Adji A. Association between Brachial-Ankle Pulse Wave Velocity as a Marker of Arterial Stiffness and Body Mass Index in a Chinese Population. J Cardiovasc Dev Dis. 2022;9. doi: 10.3390/jcdd9030075

12. Laurent S, Cockcroft J, Van Bortel L, Boutouyrie P, Giannattasio C, Hayoz D, Pannier B, Vlachopoulos C, Wilkinson I, Struijker-Boudier H, European Network for Non-invasive Investigation of Large A. Expert consensus document on arterial stiffness: methodological issues and clinical applications. Eur Heart J. 2006;27:2588–2605. doi: 10.1093/eurheartj/ehl254

13. Falcioni L, Gallotta MC, Baldari C, Cardinali L, Campanella M, Ferrari D, Guidetti L, Meucci M. Influence of training status on cardiac and vascular functioning in young recreational and competitive male rowers. Front Pediatr. 2023;11:1162594. doi: 10.3389/fped.2023.1162594

14. Perrault R, Omelchenko A, Taylor CG, Zahradka P. Establishing the interchangeability of arterial stiffness but not endothelial function parameters in healthy individuals. BMC Cardiovasc Disord. 2019;19:190. doi: 10.1186/s12872-019-1167-3

15. Boudoulas H. Systolic time intervals. Eur Heart J. 1990;11 Suppl I:93–104. doi: 10.1093/eurheartj/11.suppl_i.93

16. Reant P, Dijos M, Donal E, Mignot A, Ritter P, Bordachar P, Dos Santos P, Leclercq C, Roudaut R, Habib G, Lafitte S. Systolic time intervals as simple echocardiographic parameters of left ventricular systolic performance: correlation with ejection fraction and longitudinal two-dimensional strain. Eur J Echocardiogr. 2010;11:834–844. doi: 10.1093/ejechocard/jeq084

17. Triposkiadis F, Giamouzis G, Boudoulas KD, Karagiannis G, Skoularigis J, Boudoulas H, Parissis J. Left ventricular geometry as a major determinant of left ventricular ejection fraction: physiological considerations and clinical implications. Eur J Heart Fail. 2018;20:436–444. doi: 10.1002/ejhf.1055

18. Biering-Sorensen T, Querejeta Roca G, Hegde SM, Shah AM, Claggett B, Mosley TH, Jr., Butler KR, Jr., Solomon SD. Left ventricular ejection time is an independent predictor of incident heart failure in a community-based cohort. Eur J Heart Fail. 2018;20:1106–1114. doi: 10.1002/ejhf.928

19. Chirinos, J.A. Textbook of arterial stiffness and pulsatile hemodynamics in health and disease. Academic Press; 2022.

20. Tsiachris D, Tsioufis C, Syrseloudis D, Roussos D, Tatsis I, Dimitriadis K, Toutouzas K, Tsiamis E, Stefanadis C. Subendocardial viability ratio as an index of impaired coronary flow reserve in hypertensives without significant coronary artery stenoses. J Hum Hypertens. 2012;26:64–70. doi: 10.1038/jhh.2010.127

21. Park JS, Shin JH, Park JB, Choi DJ, Youn HJ, Park CG, Kwan J, Ahn Y, Kim DW, Rim SJ, et al. Central hemodynamics and the discrepancy between central blood pressure and brachial blood pressure. Medicine (Baltimore). 2022;101:e30484. doi: 10.1097/MD.0000000000030484

22. Aursulesei Onofrei V, Ceasovschih A, Anghel RC, Roca M, Marcu DTM, Adam CA, Mitu O, Cumpat C, Mitu F, Crisan A, et al. Subendocardial Viability Ratio Predictive Value for Cardiovascular Risk in Hypertensive Patients. Medicina (Kaunas). 2022;59. doi: 10.3390/medicina59010024

23. Ng M, Fleming T, Robinson M, Thomson B, Graetz N, Margono C, Mullany EC, Biryukov S, Abbafati C, Abera SF, et al. Global, regional, and national prevalence of overweight and obesity in children and adults during 1980-2013: a systematic analysis for the Global Burden of Disease Study 2013. Lancet. 2014;384:766–781. doi: 10.1016/S0140-6736(14)60460-8

24. Pickering TG, Hall JE, Appel LJ, Falkner BE, Graves J, Hill MN, Jones DW, Kurtz T, Sheps SG, Roccella EJ. Recommendations for blood pressure measurement in humans and experimental animals: part 1: blood pressure measurement in humans: a statement for professionals from the Subcommittee of Professional and Public Education of the American Heart Association Council on High Blood Pressure Research. Circulation. 2005;111:697–716. doi: 10.1161/01.CIR.0000154900.76284.F6

25. Butlin M, Qasem A. Large Artery Stiffness Assessment Using SphygmoCor Technology. Pulse (Basel). 2017;4:180–192. doi: 10.1159/000452448

26. Jia G, Habibi J, Aroor AR, Martinez-Lemus LA, DeMarco VG, Ramirez-Perez FI, Sun Z, Hayden MR, Meininger GA, Mueller KB, et al. Endothelial Mineralocorticoid Receptor Mediates Diet-Induced Aortic Stiffness in Females. Circ Res. 2016;118:935–943. doi: 10.1161/CIRCRESAHA.115.308269

27. Aroor AR, Jia G, Sowers JR. Cellular mechanisms underlying obesity-induced arterial stiffness. Am J Physiol Regul Integr Comp Physiol. 2018;314:R387–R398. doi: 10.1152/ajpregu.00235.2016

28. Sehgel NL, Vatner SF, Meininger GA. “Smooth Muscle Cell Stiffness Syndrome”-Revisiting the Structural Basis of Arterial Stiffness. Front Physiol. 2015;6:335. doi: 10.3389/fphys.2015.00335

29. Nosalski R, Guzik TJ. Perivascular adipose tissue inflammation in vascular disease. Br J Pharmacol. 2017;174:3496–3513. doi: 10.1111/bph.13705

30. Foote CA, Castorena-Gonzalez JA, Staiculescu MC, Clifford PS, Hill MA, Meininger GA, Martinez-Lemus LA. Brief serotonin exposure initiates arteriolar inward remodeling processes in vivo that involve transglutaminase activation and actin cytoskeleton reorganization. Am J Physiol Heart Circ Physiol. 2016;310:H188–198. doi: 10.1152/ajpheart.00666.2015

31. Ford ML, Tomlinson LA, Chapman TP, Rajkumar C, Holt SG. Aortic stiffness is independently associated with rate of renal function decline in chronic kidney disease stages 3 and 4. Hypertension. 2010;55:1110–1115. doi: 10.1161/HYPERTENSIONAHA.109.143024

32. DeMarco VG, Habibi J, Jia G, Aroor AR, Ramirez-Perez FI, Martinez-Lemus LA, Bender SB, Garro M, Hayden MR, Sun Z, et al. Low-Dose Mineralocorticoid Receptor Blockade Prevents Western Diet-Induced Arterial Stiffening in Female Mice. Hypertension. 2015;66:99–107. doi: 10.1161/HYPERTENSIONAHA.115.05674

33. Bagi Z, Broskova Z, Feher A. Obesity and coronary microvascular disease - implications for adipose tissue-mediated remote inflammatory response. Curr Vasc Pharmacol. 2014;12:453–461. doi: 10.2174/1570161112666140423221843

